# COVID-19 Health Care Behaviour in The Gambia: a cross-sectional survey of 205 adults who went through mandatory institutional quarantine

**DOI:** 10.1101/2021.11.17.21266451

**Authors:** Penda Johm, Oluwatosin Nkereuwem, Aji-Matty Manjang, Omar Ceesay, Lamin Leigh, Amie Ceesay, Mustapha Bittaye, Adeyemi Roberts, Buba Manjang, Sana Sambou, Sainey Sanneh, Lamin Saidy, Binta Saidy, Beate Kampmann

## Abstract

**Background:** To control the spread of the novel Coronavirus disease 2019 (COVID-19) caused by severe acute respiratory syndrome novel Coronavirus-2 (SARS-CoV-2), countries around the world subsequently implemented several public health measures, including the adoption of mandatory institutional quarantine for close contacts. This study explored the experiences of individuals who underwent institutional quarantine in The Gambia to inform government measures to increase its effectiveness and reduce its associated negative impacts.

**Methods:** Questionnaires were administered via mobile phone call with data collectors calling and directly recording participant responses on a tablet in an electronic online form developed in REDCap (Research Electronic Data Capture). The questionnaire contained questions on COVID-19 related knowledge, health care behaviour, attitudes, perceptions and stigma. Data were analysed using STATA v.13 (Stata Corp, College Station, TX, USA).

**Results:** In total, 205 adults who observed the mandatory institutional quarantine were interviewed. There was varied knowledge of COVID-19 causes, spread, symptoms, diagnosis, treatment, and severity. Participants believed the purpose of quarantine was monitoring for signs and symptoms of coronavirus disease, testing for SARS-CoV-2, separation from the community, and protection from coronavirus disease. While a majority reported positive experiences while in quarantine, some expressed prominent dissatisfaction related to the essential services and quality of care provided. Different forms of stigma were also experienced before, during and after the quarantine experience.

**Conclusion:** This study provides important information on quarantine experiences in The Gambia during the global COVID-19 pandemic. The Ministry of Health in The Gambia and other countries could improve the experience of quarantined individuals by consistently providing psychosocial support, compensation for loss of earnings, and timely provision of SARS-CoV-2 test results. Furthermore, stigma experiences and practices should be addressed during and after individuals stay in quarantine via the provision of psychosocial support.

## INTRODUCTION

On the 11^th^ of March 2020, the World Health Organization (WHO) declared Coronavirus disease 2019 (COVID-19) a pandemic. COVID-19 is a novel disease caused by severe acute respiratory syndrome novel Coronavirus-2 (SARS-CoV-2) (1). As of the 20^th^ of August 2021, there have been more than 212 million cases and over 4.4 million deaths recorded worldwide (2). Over 147,000 new cases were reported in the African region from the 23^rd^ to 29^th^ of August 2021 (3). In The Gambia, according to the Ministry of Health (MoH), 359th Situation Report, there were 9,470 total confirmed cases, 9049 recoveries and 301 deaths as of that same date. The Gambia received its first shipment of 36,000 doses of COVID-19 vaccines through the COVAX facility on the 3^rd^ of March 2021 with rollout of vaccinations commencing in the days thereafter. Regarding COVID-19 vaccination status, of the total national target population (≥ 18 years), 132,756 have received a single dose of Johnson & Johnson’s Janssen vaccine, 35597 received a first dose of Oxford, AstraZeneca vaccine and 4,388 a first dose of Sinopharm vaccine (4).

Several public health measures have been put in place globally to reduce the transmission of COVID-19 and to minimize the impact of the disease. This includes risk communication, surveillance, contact tracing, social/physical distancing and mandatory institutional quarantine. Contact tracing has been used to quickly determine whether any secondary cases of COVID-19 had occurred due to an infected person encountering other people. Those contacts are then given information on how to maintain preventive measures during quarantine and what to do if they develop symptoms of COVID-19, including where to be tested (5). Institutional quarantine has been used to restrict the activities of individuals who are not ill with COVID-19 but may have been in close contact with a person with signs and symptoms of COVID-19 or travelled from an area with high transmission of COVID-19, with the aim of protecting unexposed members of the community from contracting the disease (6). In The Gambia, quarantined individuals stayed isolated from others except key hotel and MoH staff at a facility identified by the government for a mandatory period of 14 days. Those held at the facilities were provided with food, shelter, and medical assistance at the expense of the government. All quarantined individuals were required to be tested twice during quarantine, and if found positive, they were transferred to an isolation centre identified by the MoH. If found negative after 14 days, they were discharged and provided with a medical certificate. A recent study in China highlights the success of institutional quarantine in minimizing the risk of community spread of COVID-19 (7).

As the pandemic continues to evolve, it is important to understand health care behaviour in different geographical and economic settings, evaluate adherence to epidemic prevention and treatment responses and disease related stigma across the world at different stages of the global response. Adhering to national COVID-19 guidance is not necessarily feasible for everyone. Those who live in overcrowded conditions may not be able to practice social distancing and those without access to clean water and soap will not be able to wash their hands frequently (8). Other individuals may not know or understand the specific steps to follow or may not be convinced of the need to practice these behaviours. Another barrier to engagement in health care and adherence to treatment for health conditions is stigma (9). Stigma is a powerful element in determining health behaviour and is one reason for social isolation and exclusion. Stigma stems from individual characteristics as well as context-specific cultural values, norms, and attitudes. Once a stigma is applied, it manifests in a range of stigma experiences and practices such as verbal abuse or gossip (10).

Taking these factors into consideration, we documented the experiences of individuals in The Gambia who completed the government mandated institutional quarantine to develop effective recommendations for the MoH in The Gambia and other similar cultural settings to better support people before, during and after quarantine. We explored their experiences and perceptions of COVID-19 behaviour change measures, including adherence to mitigation gestures such as self-isolation and social/physical distancing and factors influencing COVID-19 related stigma.

## METHODS

### Ethical considerations

This study received approval from the Gambia Government/MRC Joint Ethics Committee (Ref. 22271) on the 11^th^ of August 2020 and the London School of Hygiene & Tropical Medicine (LSHTM) Observational/Interventions Research Ethics Committee (Ref. 22271) on the 17^th^ of August 2020. Joining the study was voluntary and participants personal information remained confidential. Unique identifiers were allocated to each person prior to being sensitized and names were not recorded on study documents but only on encrypted databases. After sensitization and before consenting, data collectors asked participants if they understood everything read out to them and gave them the opportunity to ask questions and discuss answers. The survey had an introductory text stating the survey was completely anonymous, and consent was implied by respondents agreeing to participate and completing the survey. Participants also carried out an Assessment of Understanding (AoU), wherein they needed to answer at least three out of four questions correctly with 89.76% success rate at first attempt and 10.24% success rate at second attempt.

### Study setting

The Gambia is the smallest country on mainland Africa, specifically located in West Africa, and bordered by Senegal on three sides. The nation has two main religions and eight main ethnic groups (11). According to The Gambia Multiple Indicator Cluster Survey (MICS) 2018 (12), the percentage of women and men aged 15-24 years who were able to read a short simple statement about everyday life or who attended secondary or higher education was 64.3 and 68.0 respectively. 74% of women and 85% of men aged 15-49 years owned a mobile phone. In the latest Gambia Demographic and Health Survey (GDHS) 2019-20 (13), a total of 11,865 women and 4,636 men were interviewed, representing a response rate of 95% female and 87% male respectively. 47% of women and 67% of men aged 15-49 were literate. Only 2% of women and 7% of men accessed the newspaper, television, and radio on a weekly basis. These indicators affect how Gambian people respond in a pandemic such as COVID-19 that requires knowledge and effective communication.

### Study design and population

We conducted a cross-sectional mobile phone survey from April to September 2020 to capture quantitative data from a list of individuals who had been institutionally quarantined. This password-protected person list and contact details were obtained through the MoH with approval from the Director of Health Services. The list included the name, arrival date, gender, age, address, phone number, room number, nationality, country and date of departure of every individual known to had come into The Gambia by air or land travel and through contact tracing. During this time, a total of 863 individuals were put in compulsory institutional quarantine in accordance with the government regulation in The Gambia, accommodated in four hotels usually housing tourists and based at the coast. We filtered the list based on study inclusion criteria and attempted to reach every single individual through phone calls.

### Criteria for sample selection

We enrolled all individuals who completed compulsory quarantine for COVID-19 in The Gambia, aged 18 or above, residing in The Gambia, with a valid phone number, and documented consent to take part in the study.

### Data collection

Questionnaires were administered via mobile phone calls with participant responses recorded on a tablet in an electronic online form developed in REDCap (Research Electronic Data Capture) and hosted at the Medical Research Council Unit The Gambia at the London School of Hygiene and Tropical Medicine (MRCG at LSHTM) in the Gambia. Data collectors (5 males, 4 females) with experience conducting surveys in The Gambia and fluent in English and at least one local language called every person fitting inclusion criteria to sensitize and recruit them, noting those who refused to participate or were unreachable. During this initial call, a summary of the study information sheet in either English, Mandinka, Wolof, or Fula were read out and the full study information sheet was sent to retain via WhatsApp. The study interviewer would re-attempt the phone call two more times on two separate days if initially unreachable. A third unsuccessful attempt was noted down as a non-response. At the time of data collection, all persons had exited the quarantine centres. Questionnaire answers were collected remotely with a pretested questionnaire to avoid any face-to-face contact due to the rising cases of COVID-19 at the time. The survey was pre-tested and included questions covering individual knowledge, attitudes, and perceptions of their quarantine experience. Demographic characteristics were also captured including sex, age, and highest level of education.

### Variables and data analysis

The REDCap data dictionary codebook contained a total of 76 survey questions/fields with items of different formats; closed-ended (multiple choice, dichotomous, likert scale, self-assessment with an alternative titled “other”) and open-ended to obtain in-depth information on opinions and attitudes (Supplementary File 1). “Other” responses were evaluated and reclassified into one of the existing or created new categories. Data were analysed using STATA v.13 (Stata Corp, College Station, Texas, USA) for descriptive summary statistics (frequencies and percentages). Responses to the open-ended questions were analysed thematically using an inductive framework approach. This involved reading and re-reading the dataset to develop familiarity of the data. We then iteratively developed coding and themes that identified important elements related to the survey questions (14, 15). Initial themes were determined based on the prevalence of recurring responses across the dataset and from there the overarching themes and corresponding sub-themes for each question were identified.

### Theoretical frameworks

The frameworks of health seeking behaviour (HSB) and stigma informed the development of our questionnaire. Behaviour is a complex phenomenon, influenced by factors within the individual as well as without (family and peer networks, community, and society). The Partners for Applied Social Sciences (PASS) model describes the path people follow when seeking care and focuses on the factors involved in each step that hinder or facilitate access to care (16). The Health Stigma and Discrimination Framework shows the stigmatization process as it unfolds across the social ecological spectrum, including drivers and facilitators such as race, class, gender, sexual orientation, or occupation. Once a stigma is applied, it manifests in a range of stigma experiences and practices. Stigma experiences can include experienced discrimination, which refers to stigmatizing behaviours that fall within the purview of the law, or stigmatizing behaviours that fall outside the purview of the law such as verbal abuse or gossip. Another experience of stigma is self-stigma, wherein a stigmatized person adopts society’s negative beliefs and feelings regarding their status (10). These frameworks were selected as they are not only theoretical but facilitate an understanding of the factors that enable health care behaviour, adherence, and stigma.

## RESULTS

### Participant characteristics

Out of the 429 individuals eligible, 201 were categorized as non-response (123 switched off, 51 no answer, 13 wrong numbers, 12 unavailable, 1 died, 1 one mentally unstable), 21 refused, 2 withdrew and 228 were sensitized. A total of 205 adults accepted and were interviewed between September to December 2020 (Figure 1). Participants were mostly males (81.1%), Gambian nationals (84.4%) and Muslims (92.7%). The socio-demographic characteristics of participants are shown in Table 1.

**Table 1:**
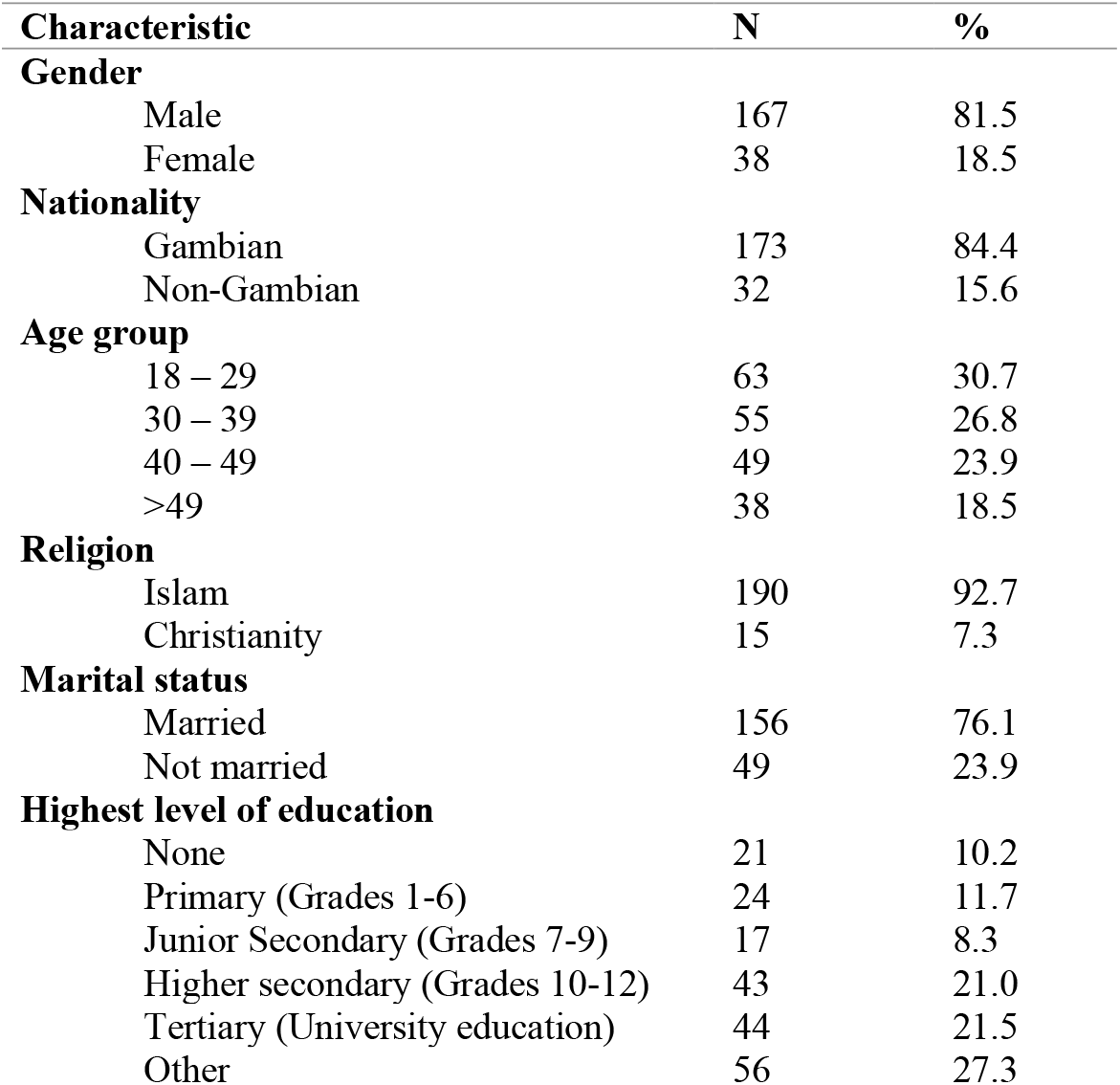
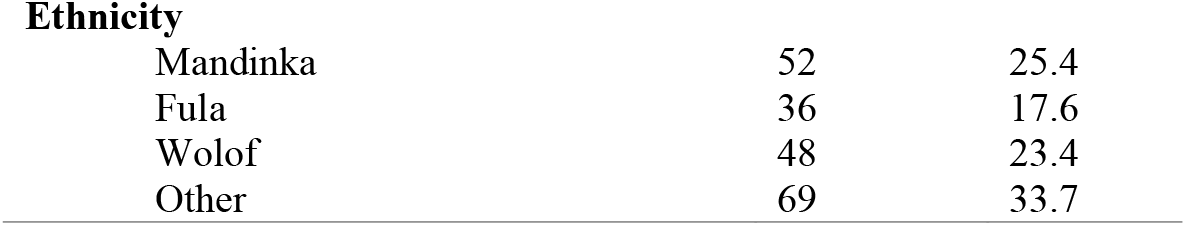
Socio-Demographic Characteristics of Participants (N=205)

| Characteristic | N | % |
| --- | --- | --- |
| **Gender** |  |  |
| Male | 167 | 81.5 |
| Female | 38 | 18.5 |
| **Nationality** |  |  |
| Gambian | 173 | 84.4 |
| Non-Gambian | 32 | 15.6 |
| **Age group** |  |  |
| 18 – 29 | 63 | 30.7 |
| 30 – 39 | 55 | 26.8 |
| 40 – 49 | 49 | 23.9 |
| >49 | 38 | 18.5 |
| **Religion** |  |  |
| Islam | 190 | 92.7 |
| Christianity | 15 | 7.3 |
| **Marital status** |  |  |
| Married | 156 | 76.1 |
| Not married | 49 | 23.9 |
| **Highest level of education** |  |  |
| None | 21 | 10.2 |
| Primary (Grades 1-6) | 24 | 11.7 |
| Junior Secondary (Grades 7-9) | 17 | 8.3 |
| Higher secondary (Grades 10-12) | 43 | 21.0 |
| Tertiary (University education) | 44 | 21.5 |
| Other | 56 | 27.3 |
| **Ethnicity** |  |  |
| Mandinka | 52 | 25.4 |
| Fula | 36 | 17.6 |
| Wolof | 48 | 23.4 |
| Other | 69 | 33.7 |

**Figure 1.**
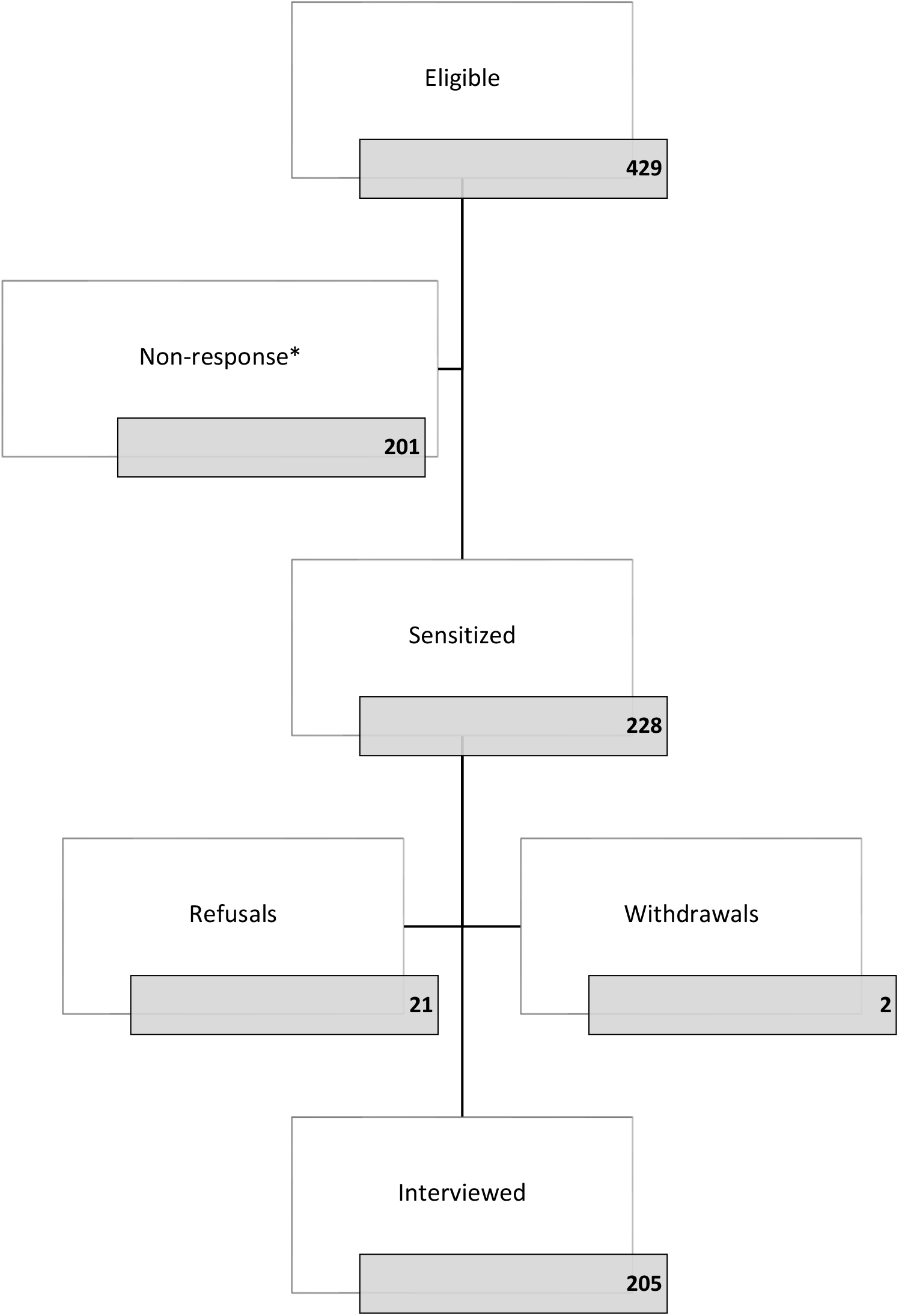
Participant recruitment process based on study inclusion criteria. *Switched off 123; No answer 51; Wrong number 5; Busy/unavailable 12; Died 1; Mentally unstable 1; Wrong person 8

### Knowledge and perceptions of coronavirus disease 2019, COVID-19

The initial questions probed our participants knowledge base (Table 2). Social and mass media were identified as the most reported first source of information regarding COVID-19 and almost all participants trusted this first source of information. When asked about the mode of acquiring COVID-19 was, nearly half of participants mentioned droplets from infected people when they cough or sneeze. About a quarter said direct contact with an infected person. Almost half of our participants identified fever and coughing as symptoms of COVID-19. Most of our participants said they believed that COVID-19 was deadly and about half identified elderly people as the most at-risk population of being infected with the disease. Almost all said they worried that they were at risk of contracting coronavirus disease.

**Table 2:** Knowledge and Perceptions of Coronavirus Disease 2019, COVID-19 (N=205)

| Variable | Category | N | % |
| --- | --- | --- | --- |
| **From whom or from where did you FIRST hear about coronavirus disease, also known as COVID-19? (choose one)** | Family member | 11 | 5.4 |
|  | Health staff/Health centre | 14 | 6.8 |
|  | Mass media/Social media | 158 | 77.1 |
|  | Religious centre | 2 | 1.0 |
|  | Workplace | 4 | 2.0 |
|  | Other | 16 | 7.8 |
| **Did you trust the source from which you FIRST heard about coronavirus disease?** | Yes | 168 | 82.0 |
|  | No | 37 | 18.0 |
| **How does coronavirus disease spread/how can someone get infected with coronavirus disease? (multiple choice)** | Don't know | 11 | 2.0 |
|  | Droplets from infected people when they cough or sneeze | 265 | 49.1 |
|  | Direct contact with an infected person | 143 | 26.5 |
|  | Touching objects/surfaces contaminated by an infected person | 102 | 18.9 |
|  | Other (please specify) | 19 | 3.5 |
| **What are the symptoms of coronavirus disease, COVID-19? (multiple choice)** | Don't know | 14 | 1.9 |
|  | Fever | 160 | 21.6 |
|  | Sore throat | 90 | 12.1 |
|  | Coughing | 156 | 21.1 |
|  | Breathing difficulties | 95 | 12.8 |
|  | Body pains | 66 | 8.9 |
|  | Headache | 78 | 10.5 |
|  | Loss of smell | 26 | 3.5 |
|  | Loss of taste | 30 | 4.0 |
|  | Other | 26 | 3.5 |
| **Do you believe that coronavirus disease 2019, COVID-19, is deadly?** | Yes | 181 | 88.3 |
|  | No | 24 | 11.7 |
| **Who do you think is most at risk of being infected with coronavirus disease? (choose one)** | Don't know | 8 | 3.9 |
|  | Elderly | 103 | 50.2 |
|  | Persons with underlying conditions | 27 | 13.2 |
|  | Those that did not follow guidelines | 5 | 2.4 |
|  | Health workers | 6 | 2.9 |
|  | Everyone | 44 | 21.5 |
|  | Other | 12 | 5.9 |
| **Do you worry that you are at risk of contracting coronavirus disease?** | Yes | 162 | 79.0 |
|  | No | 43 | 21.0 |
| **What do you consider important actions to prevent the spread of coronavirus disease in your community? (multiple choice)** | Staying at home | 48 | 7.95 |
|  | Washing hands regularly using hand soap and water or hand sanitizer | 136 | 22.52 |
|  | Testing people for coronavirus disease | 52 | 8.61 |
|  | Community sensitization | 107 | 17.72 |
|  | Everyone wearing a mask when out in public | 111 | 18.38 |
|  | Social/physical distancing | 112 | 18.54 |
|  | People get vaccinated against coronavirus disease | 15 | 2.48 |
|  | Other | 23 | 3.81 |

### COVID-19 Health Care Behaviour

We assessed the way participants had sought health care for themselves before and during quarantine (Table 3). Most of our participants came to be quarantined following land travel into the country or the MoH contact tracing. When asked what they understood to be the purpose of the quarantine process, a majority mentioned monitoring for signs and symptoms of COVID-19, with some saying they were there to be tested for COVID-19. As we further explored the health care behaviour of our consenting participants, almost half reported that they would visit a hospital or health facility as their first point of care if they experienced symptoms of COVID-19. Exploring adherence, participants were asked which quarantine measures they did or did not comply with during their stay at the quarantine centre. Almost all (99%) reported that they stayed at their place of quarantine every day. 66% did not allow any visitors compared to 34% who did. The majority separated themselves from others at the place where they were being quarantined (93%), did not share food and utensils with others (81%), and wore a face mask if they had to leave their room (76%).

**Table 3:**
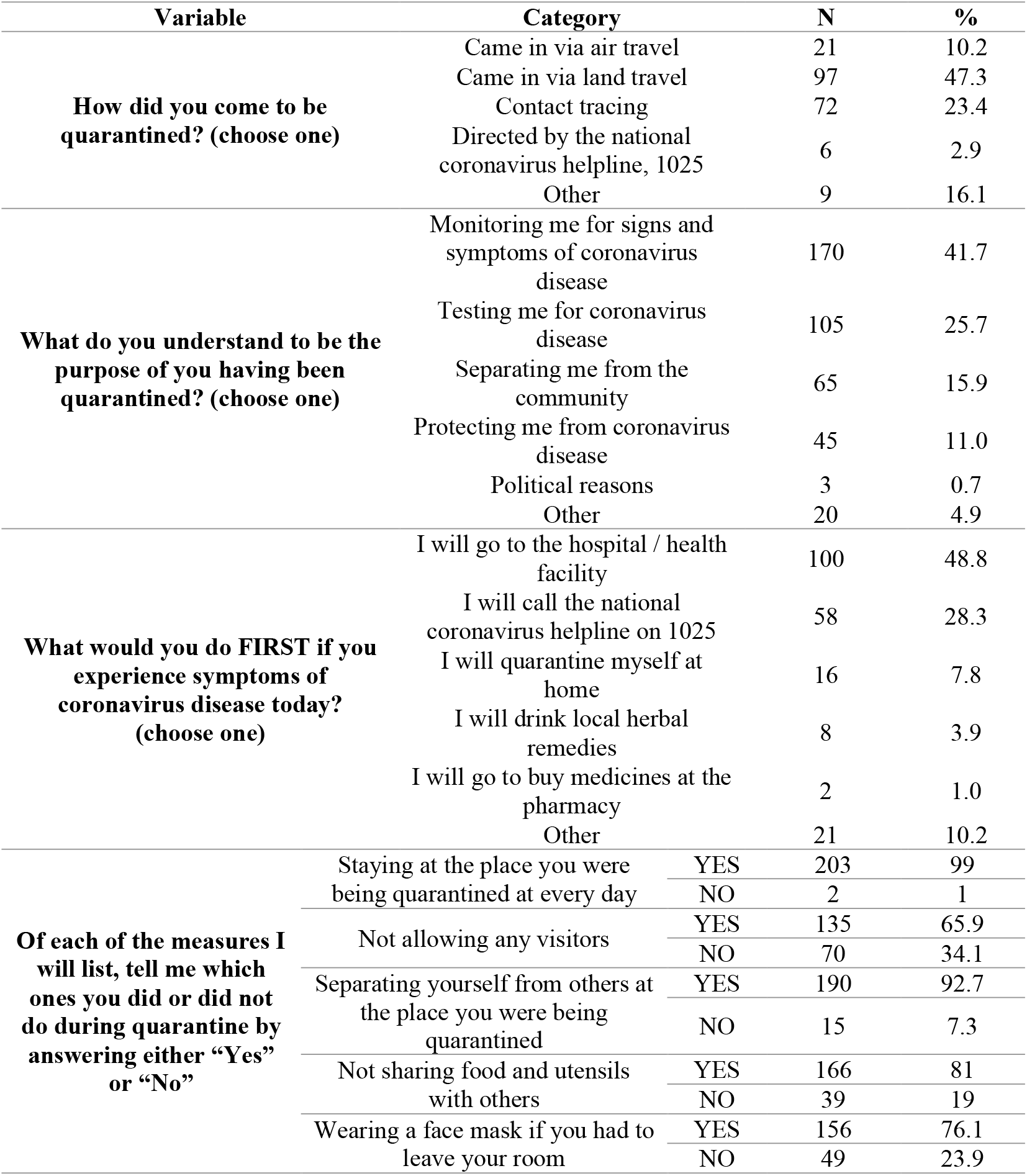
COVID-19 Health Care Behaviour (N=205)

### COVID-19 Related Stigma

Participants were asked questions about stigma regarding their experience before, during and after going into quarantine (Table 4). COVID-19 related stigma was hardly experienced during quarantine. However, a significant number (40%) had people suggesting they were to go into quarantine prior to being admitted. More experienced stigma after quarantine, with people gossiping about them (38%), family and friends refusing to visit them at home (19.5), anyone who called thinking they had been treated for coronavirus disease (27.8%), and even being called names to make them feel ashamed (25.4). Participants also experienced enacted stigma (overt discrimination against the stigmatized) after quarantine leading to feelings of shame, self-blame, loneliness, stress, anger and withdrawal from daily activities.

**Table 4:**
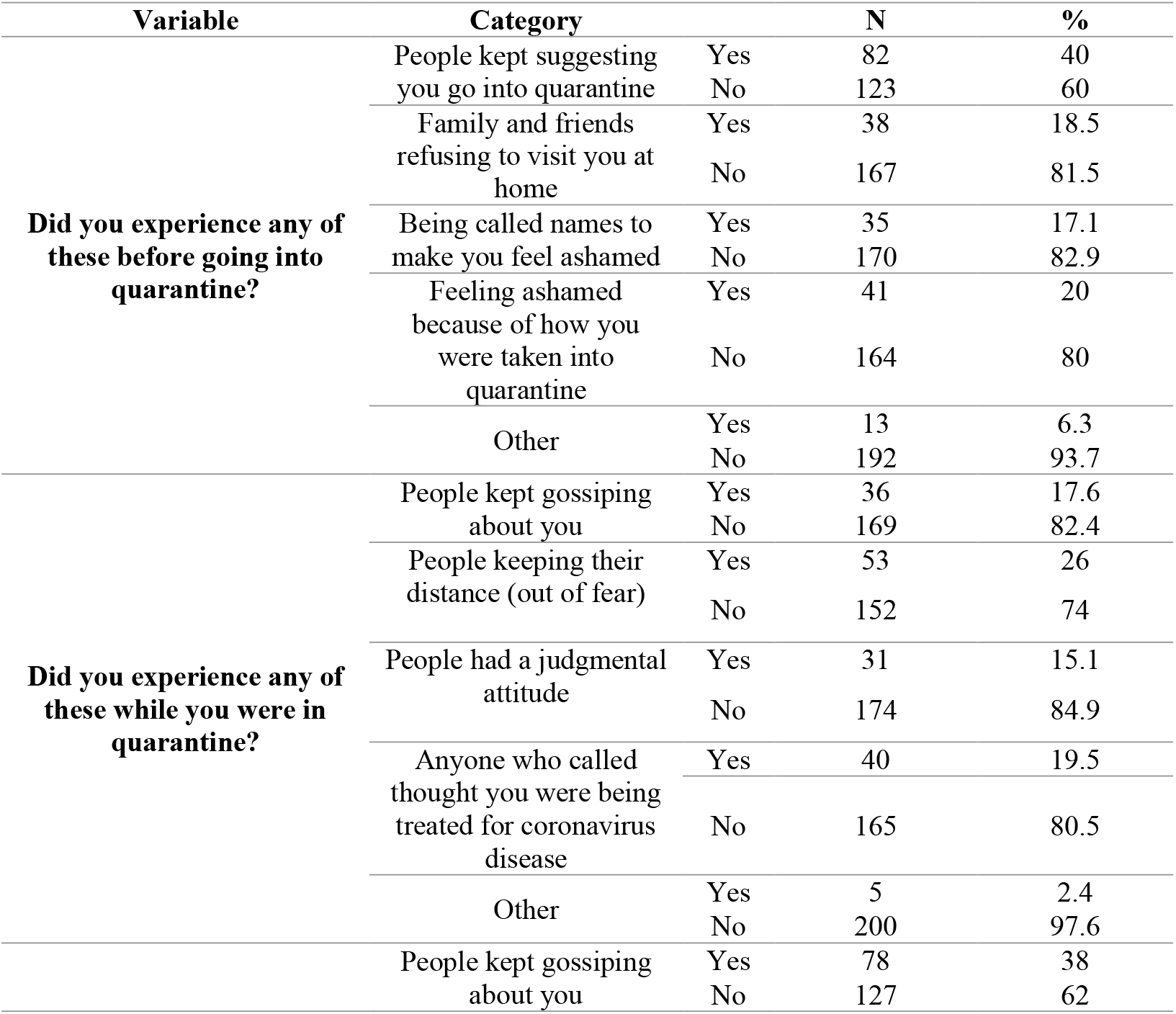
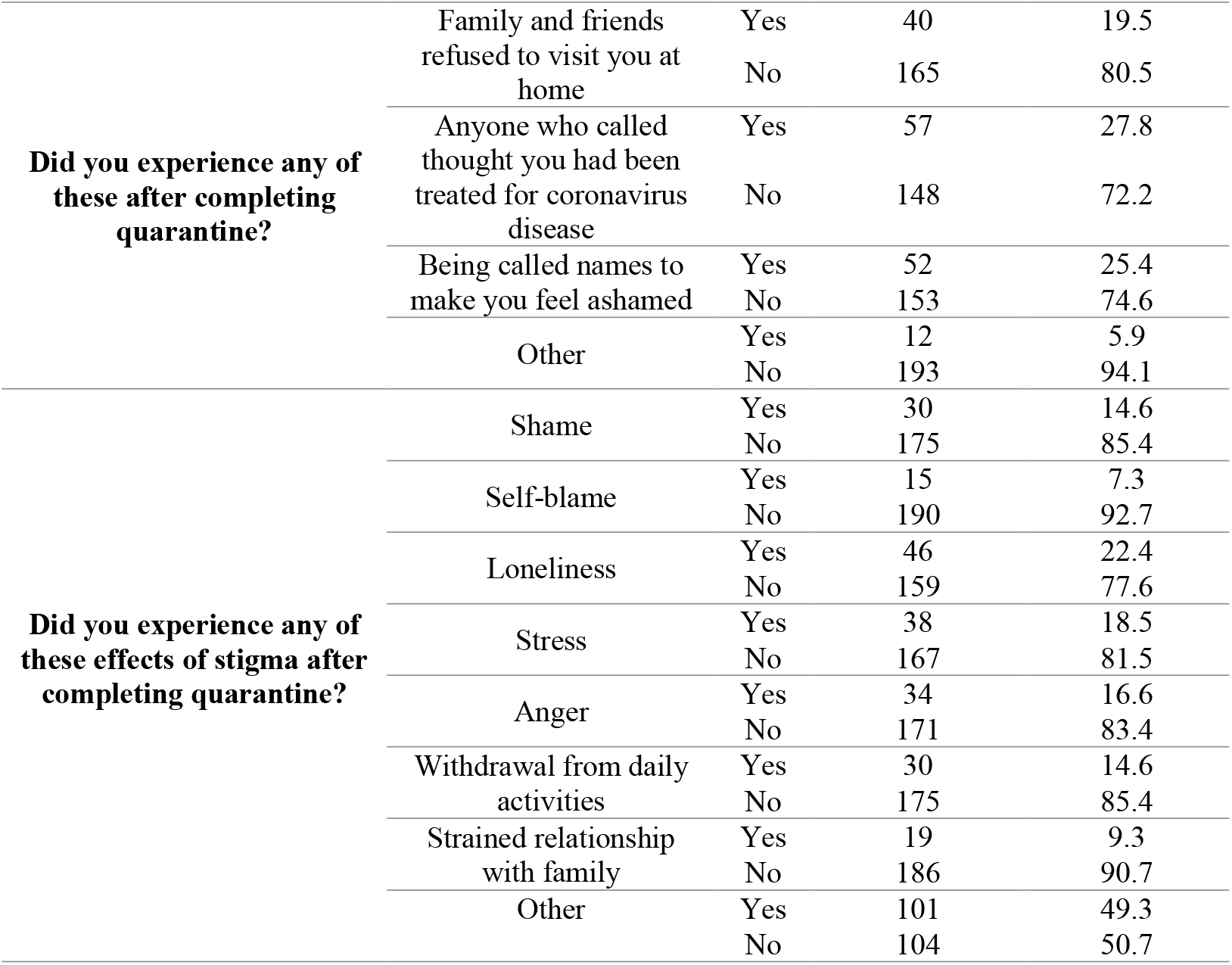
COVID-19 Related Stigma (N=205)

Attitudes towards COVID-19 related stigma were measured using a Likert scale, wherein respondents rated their levels of agreement with different statements (Figure 2). Over 80% strongly agreed or agreed that judging and shaming is hurtful and can negatively affect mental health. Almost 90% strongly agreed or agreed that recovering from coronavirus disease is nothing to be ashamed about or blamed for. Over 80% strongly agreed or agreed that people may refuse to go into quarantine for fear of being accused of having coronavirus disease. There was discrepancy regarding whether public judgement or shaming of others is caused by the publics fear of being infected with coronavirus disease. 70% strongly agreed or agreed whereas 28% disagreed. There was also discrepancy regarding whether people who recover from coronavirus disease will be shamed and blamed for having it. More than half strongly disagreed or disagreed compared to over 40% who strongly agreed or agreed.

**Figure 2.**
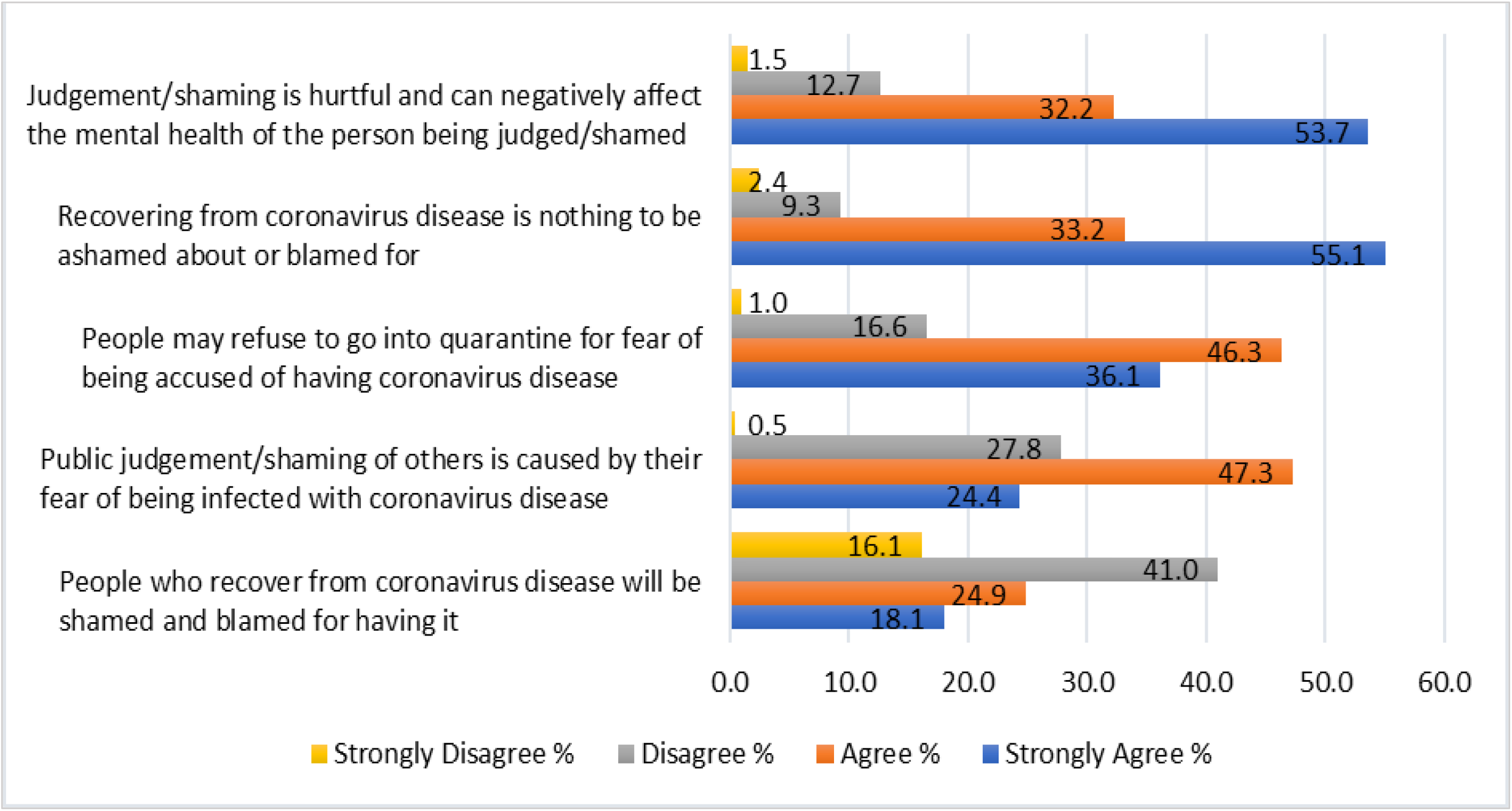
Likert scale gauging attitudes towards COVID-19 related stigma.

### Perceptions of quarantine experience

Participants were also asked to evaluate their quarantine experience (Table 5). Most participants (68.29%) shared positive quarantine experiences in the facilities where they were accommodated. While some considered their stay a good experience, some said they had time to relax after many years of working without rest, others had time for spiritual activities, and most commended the efforts of the facility staff who they considered friendly. Almost a third (29.27%) of the participants expressed negative experiences, some participants were upset while sharing their experiences with our team. Listed benefits of quarantine were testing for and knowing their COVID-19 status; separating potentially positive cases from others in the community; reading the Quran and having more time to worship God; care provided by health workers and hotel staff; relaxation; adequate food and housing; and receiving a certificate after completion of quarantine. Disadvantages were cited as being away from and missing family; not being able to provide financially for the family as the household head; poor taste and limited quantity of food served at the hotel; loneliness, lack of socialisation and social activities; no receipt of COVID-19 test results; lots of mosquitoes and no bed nets; and time wasted with no remuneration.

**Table 5:**
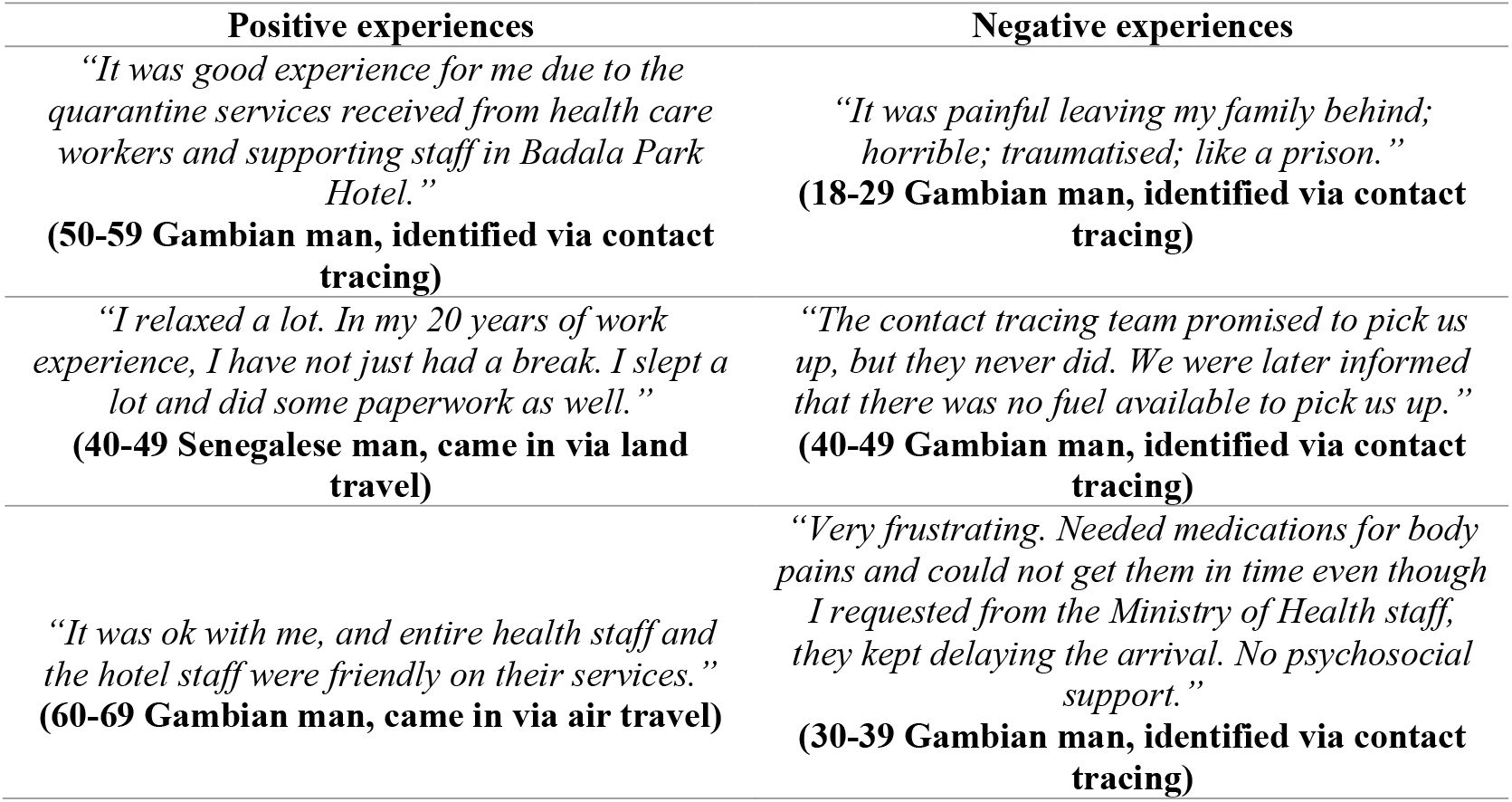
Perceptions of Quarantine Experience.

| Positive experiences | Negative experiences |
| --- | --- |
| *“It was good experience for me due to the quarantine services received from health care workers and supporting staff in Badala Park Hotel.”*<br>**(50-59 Gambian man, identified via contact tracing)** | *“It was painful leaving my family behind; horrible; traumatised; like a prison.”*<br>**(18-29 Gambian man, identified via contact tracing)** |
| *“I relaxed a lot. In my 20 years of work experience, I have not just had a break. I slept a lot and did some paperwork as well.”*<br>**(40-49 Senegalese man, came in via land travel)** | *“The contact tracing team promised to pick us up, but they never did. We were later informed that there was no fuel available to pick us up.”*<br>**(40-49 Gambian man, identified via contact tracing)** |
| *“It was ok with me, and entire health staff and the hotel staff were friendly on their services.”*<br>**(60-69 Gambian man, came in via air travel)** | *“Very frustrating. Needed medications for body pains and could not get them in time even though I requested from the Ministry of Health staff, they kept delaying the arrival. No psychosocial support.”*<br>**(30-39 Gambian man, identified via contact tracing)** |

## DISCUSSION

As a response to the COVID pandemic, many nations implemented mandatory institutional quarantine with the aim of protecting unexposed individuals from contracting the disease, but there is little insight into the experiences of individuals who had been through these procedures. Our interview-based study explored the topics of COVID-19 knowledge, risk perceptions, attitudes, adherence, and stigma amongst individuals who had been quarantined in The Gambia to inform governmental practices that could increase effectiveness of such pandemic control measures and reduce associated negative impacts.

While exploring knowledge, we found considerable similarities to data from a recent online survey on the knowledge, attitude, and practice of Sudanese people towards COVID-19 (17), with top listed sources of knowledge acquisition of COVID-19 being mass media and social media. Predominantly listed COVID-19 related symptoms identified by our participants such as fever, coughing, breathing difficulties and sore throat were consistent with widespread information circulating at the time. Over 60% of participants identified droplets from coughs and sneezes as sources of infection. This finding is in line with a recently published study conducted in Ethiopia on community COVID-19 risk perception and health-seeking behaviour (18), wherein 60.3% respondents said COVID-19 could be transmitted via inhalation of a droplet from infected people and 58.0% reported airborne as a transmission route for COVID-19. Regarding perceived severity of COVID-19, 88.3% of our participants believed that coronavirus disease was deadly, like a recent study in Ethiopia (19) and in Sudan where 85.6 % and 89.5% respectively understood COVID-19 to be a dangerous disease (17).

To nudge the public towards positive epidemic prevention behaviours, individuals must perceive their susceptibility to and the severity of COVID-19 (20). Risk perceptions give us important insight into how individuals view personal risks, which in turn is an important determinant of their behaviour and adherence to protective measures (21, 22). When gauging personal risk perceptions, it was evident that 79% of our respondents worried that they were at risk of contracting coronavirus disease, similar to 81% in a recent study in Ethiopia (18). Exactly half of our participants said the elderly were most at risk of being infected with coronavirus disease, followed by everyone else, again like a study in Sudan wherein 81.7% said those at risk of developing a severe form of COVID-19 were the elderly, followed by everyone else (36%) (17). In contrast, in another study in Ethiopia examining myths, beliefs, and perceptions about COVID-19, 45.1% and 62.2% of respondents perceived that children and youth, respectively, are at a moderate risk of COVID-19 (23). This variation may be due to the difference sources and timely access of risk information of respondents.

Several studies have explored the knowledge and adoption of actions considered important to prevent the spread of coronavirus disease in the general population, (17, 23-25), however, none of these explored the perceptions of quarantined individuals. Quarantine was not listed as a preventive measure by our study participants, however, most of our participants identified monitoring for signs and symptoms of coronavirus disease as the purpose of the mandatory institutional quarantine. Regarding pathway to seeking care for COVID-19 related symptoms in the future, 48.8% and 28.3% of our study participants claimed they would either first visit a hospital or health facility or call the national coronavirus helpline on 1025, respectively, if they experienced symptoms of coronavirus disease.

Looking at facility related factors during quarantine, there was prominent dissatisfaction reported by most of our participants regarding the facility environment, specifically cleanliness and the services provided, specifically meals. Our findings are similar to the experiences of persons in COVID-19 institutional quarantine in Uganda (26). In The Gambia, all quarantine related costs were covered by the government and therefore our study participants were not financially burdened with the cost of accommodation, food, testing and certification. Nevertheless, a predominant dissatisfaction related to cost noted by our study participants was specifically the desire to have been provided with compensation in the form of cash, food and/or transport for their time spent in quarantine, which would have benefited their families. In other countries, support measures such as financial support, employment benefits and/or practical support were adopted to enable people to follow self-isolation or quarantine guidance. In Germany, all employed people in mandatory self-isolation who tested positive were provided with 100% remuneration of their salary for up to six weeks and in South Korea, any person required to quarantine for 14 days was provided with daily necessities, sanitary kits and financial support (27). Although these findings are from high income countries, similar measures could be adapted in low- and middle-income countries such as The Gambia.

Adherence to COVID-19 prevention and control measures for quarantined individuals are key to the success of institutional quarantine. Most participants claimed to have adhered to the measures, however, few deviated and disregarded guidelines. From the limited literature published globally on adherence to self-isolation measures, it has been found that public trust, an altruistic attitude, and access to information increase adherence to social/physical distancing protocols (27, 28). We utilized a conceptual framework for quarantine acceptance and adherence to investigate variables affecting adherence to mandatory COVID-19 quarantine (29). In relation to the framework, our results showed that individuals life circumstances and quarantine measures being enforced were the main predictors of their adherence.

Stigma experiences such as discrimination and behaviours such gossip can negatively affect the mental health of the stigmatized individual and lead to social exclusion. Stigma practices can include stereotypes, prejudice, stigmatizing behaviours, and discriminatory attitudes (10). In this study, COVID-19 related stigma was hardly experienced before and during quarantine but rather following on from quarantine. Some participants reported people gossiping about them, family and friends refusing to visit them at home and even being called names to make them feel ashamed. Others had people call them thinking they were being treated for coronavirus disease at the quarantine facility, and misunderstandings such as these stem from public fears of being infected by people in quarantine (7). Our participants also experienced overt discrimination (enacted stigma) after quarantine leading to feelings of shame, self-blame, loneliness, stress, anger, and withdrawal from daily activities (felt stigma). Unfortunately, this is a reality many other individuals in self-isolation and quarantine have faced worldwide as found in recent studies (30, 31). It is therefore important to continually uncover and mitigate the many drivers of stigma to improve the lives of quarantined individuals.

### Study strengths and limitations

One of the key strengths of this study was carrying out interviews via mobile phone call guided by our REDCap questionnaire displayed on tablets. This method seemed most appropriate over traditional face-to-face interviewing due to the rise of new cases at the time and government COVID-19 preventive mandates such as social/physical distancing. Our chosen method ensured we generated rapid first-hand evidence and helped us save on travel costs. Another key strength of our study was conducting interviews soon after participants left quarantine to minimise their recall bias after changing environment.

It is important to note that our study population does not represent the diverse demographics of The Gambia and not every person who went through institutional quarantine was included. Most of our participants were Gambian males who are more likely to travel than women during the pandemic and therefore were more frequently identified for institutional quarantine. Our chosen methods of phone call surveys reduced our opportunity to take note of participants’ nonverbal communication. We did not explore the stigma experience of associative stigma, meaning any stigma experienced by family or friends of those who went through institutional quarantine as they were not the cohort of focus in our study.

## CONCLUSION

This study sheds light on the lives of those who had been in institutional quarantine in The Gambia and provides evidence that can be used to inform the Gambia MoH, policy makers and other key stakeholders working on epidemic preparedness and response. Such information was missing in the literature in general at the time this study was conducted. To improve the experience of individuals in designated institutional quarantine facilities, there should be daily provision of quality food and healthcare, timely provision of SARS-CoV-2 test results and negligible or complimentary associated costs of quarantine. Furthermore, stigma experiences and practices should be addressed during and after individuals stay in quarantine via the provision of psychosocial support.

## Data Availability

The datasets used and/or analysed during the current study are available from the corresponding author on reasonable request.

## ACKNOWLEDGMENTS

We express heartfelt gratitude to our study participants for their valuable time and genuine responses to our mobile phone surveys. We would like to sincerely thank the Ministry of Health staff who supported our data collection: Baboucarr Jallow, Modou Jallow, and Ebrima Keita.

## AUTHORS’ CONTRIBUTIONS

PJ conceived and designed the study with feedback from ON, AMM, OC, LL, AC, BK, MB, BM, SS and SS. AMM, OC, LL, AC, ON, BJ, MJ, EK, PJ collected data. AMM, PJ along with LS resolved data queries and cleaned the dataset. PJ and ON led data analysis and the literature search, wrote the first draft of the manuscript, and updated subsequent versions of the paper with co-author comments and inputs. PJ, ON, BK, AMM, OC, LL, AC contributed to the interpretation of results, revised the manuscript, suggested policy implications from the findings and edited the final draft. All authors read and approved the final manuscript.

## DECLARATIONS

### Consent for publication

Not applicable.

### Competing interests

None declared.

#### LIST OF ABBREVIATIONS

AoU: Assessment of Understanding
COVID-19: Coronavirus disease 2019
GDHS: Gambia Demographic and Health Survey
HSB: Health seeking behaviour
MICS: Multiple Indicator Cluster Survey
MoH: Ministry of Health
MRCG at LSHTM: Medical Research Council Unit The Gambia at the London School of Hygiene and Tropical Medicine
PASS: Partners for Applied Social Sciences
REDCap: Research Electronic Data Capture
SARS-Cov-2: Severe Acute Respiratory Syndrome Novel Coronavirus-2
WHO: World Health Organization

