## Supplemental Table 1 for "COVID-19 Health Care Behaviour in The Gambia: a cross-sectional survey of 205 adults who went through mandatory institutional quarantine"

*Supplemental Table 1: COVID-19 Related Stigma (N=205)*

| Variable | Category | N | % |
|---|---|---|---|
| Which groups of people do you think the coronavirus disease is generating judgement/unfairness/shame/blame against in your community? (multiple choice) | Anyone who called the national coronavirus helpline 1025 | 67 | 14.6 |
| | Anyone who is seen coughing or sneezing in public | 61 | 13.3 |
| | Anyone who recovered from coronavirus disease and returned to the community | 74 | 16.1 |
| | Family members of people who recovered from coronavirus disease | 47 | 10.2 |
| | Anyone who was put into quarantine by the government and returned to the community | 79 | 17.2 |
| | Family members of people who were quarantined by the government | 36 | 7.8 |
| | Anyone who came into The Gambia by air or land | 46 | 10.0 |
| | Family members of people came into The Gambia by air or land | 22 | 4.8 |
| | Other | 27 | 5.9 |
| What are your feelings towards people diagnosed with coronavirus disease? (multiple choice) | They are unfortunate people (bad luck) | 11 | 3.6 |
| | They brought the disease upon themselves by travelling during a pandemic | 6 | 2.0 |
| | They should be isolated from their family, friends and community until fully recovered | 49 | 16.1 |
| | They are people like everyone else who just happened to contract a disease | 61 | 20.0 |
| | Their condition was meant to happen (divine will of God) | 121 | 39.7 |
| | They did something bad (sinful or immoral behaviour) that is why they have the disease | 5 | 1.6 |
| | They should not be allowed to resume work until fully recovered | 12 | 3.9 |
| | Other | 40 | 13.1 |
