## Supplementary File 1 for "COVID-19 Health Care Behaviour in The Gambia: a cross-sectional survey of 205 adults who went through mandatory institutional quarantine"

### COVID-19 HCB GAMBIA

Please complete the survey below.

Thank you!

---

q1 hcare 1) Participant Identification Number

________________________________

---

**Assessment of Understanding (AoU)**

**I would now like to assess your understanding of the information I have just provided to you about the study. Please answer whether the following statements I will read out to you are either True (T) or False (F). To continue with this phone interview/survey, you will need to answer at least three out of four questions correctly. In case your first attempt is unsuccessful, I will re-read the study summary again and you will have one re-attempt to answer the questions. In case your second attempt is unsuccessful, you will unfortunately not be able to take part in the study. Do you understand? Any questions**

---

2) Date of Assessment:

________________________________

---

3) Time of Assessment

________________________________

---

4) Attempts

- ◯ 1st Attempt
- ◯ 2nd Attempt

---

5) This study involves asking what you know of coronavirus disease 2019, COVID-19

◯ True ◯ False

---

6) We will need to conduct face to face interviews with you

◯ True ◯ False

---

7) You will be given a code number instead of your name being used anywhere in the study

◯ True ◯ False

---

8) Your participation in this study is voluntary and you are free to withdraw at any time

◯ True ◯ False

---

9) Did the participant meet all the criteria

◯ Yes ◯ No

---

10) By clicking Accept, you confirm that you have read and agreed to the the Participant information sheet and Consent form.

◯ Accept ◯ Not Accept

---

11) Gender

- ◯ Male
- ◯ Female
- ◯ Other

11a) Specify Others

__________________________________

12) Date

__________________________________
((DD/MM/YYYY) )

13) Start time

__________________________________

14) Interviewer ID

__________________________________

15) Interview language

◯ English
◯ Mandinka
◯ Fula
◯ Wolof
◯ Other
(Choose One)

15a) Specify Others

__________________________________

**PART 1: Knowledge and perceptions of coronavirus disease, COVID-19**

**I would first like to ask you some questions on what you may know about coronavirus disease**

16) From whom or from where did you FIRST hear about coronavirus disease, also known as COVID-19?

◯ Family member
◯ Health staff including community health worker
◯ Health centre or hospital
◯ Mass media (Radio, TV)
◯ WhatsApp/ Social media
◯ Mosque or Church
◯ Religious leader
◯ Other
(Choose One)

16a) Specify Others

__________________________________

17) Do you trust the source from which you FIRST heard about coronavirus disease?

◯ Yes ◯ No

18) What do you know about coronavirus disease, COVID-19?

__________________________________

19) How does coronavirus disease spread/how can someone get infected with coronavirus disease?

☐ Don't know
☐ Droplets from infected people when they cough
☐ Droplets from infected people when they sneeze
☐ Direct contact with an infected person
☐ Touching objects/surfaces contaminated by an infected person
☐ Other
(Multiple Choice)

19a) Specify Others

__________________________________

| | |
|---|---|
| 20) What are the main symptoms of the coronavirus disease | ☐ Don't know<br>☐ Fever<br>☐ Sore throat<br>☐ Coughing<br>☐ Breathing difficulties<br>☐ Body pains<br>☐ Headache<br>☐ Loss of smell<br>☐ Loss of taste<br>☐ Other<br>(Multiple Choice) |
| 20a) Specify Other | __________________________<br>(e.g. vomiting, diarrhoea, etc) |
| 21) Do you believe that coronavirus disease 2019, COVID-19, is deadly? | ◯ Yes<br>◯ No |
| 22) Who do you think is most at risk of being infected with coronavirus disease? | ◯ Don't know<br>◯ Women<br>◯ Men<br>◯ Children<br>◯ Elderly<br>◯ Everyone<br>◯ Other<br>(Choose One) |
| 22a) Specify Others | __________________________ |
| 23) Do you worry that you are at risk of contracting coronavirus disease? | ◯ Yes<br>◯ No |
| 23a) Yes. Why? | __________________________ |
| 23b) No. Why not? | __________________________ |
| 24) Do you know how to prevent becoming infected with coronavirus | ◯ Yes<br>◯ No<br>(If no, skip to Q26) |
| 25) What measures have you taken to prevent becoming infected with coronavirus disease in recent days? | ☐ Staying at home<br>☐ Washing hands regularly using hand soap and water or hand sanitizer<br>☐ Covering your mouth and nose when coughing or sneezing<br>☐ Avoiding close contact with anyone who has a fever and/or cough<br>☐ Other<br>(Multiple Choice) |
| 25a) Specify Others | __________________________ |

26) What do you consider important actions to prevent the spread of coronavirus disease in your community

- ☐ Staying at home
- ☐ Washing hands regularly using hand soap and water or hand sanitizer
- ☐ Testing people for coronavirus disease
- ☐ Community sensitization
- ☐ Everyone wearing a mask when out in public
- ☐ Social/physical distancing
- ☐ People get vaccinated against coronavirus disease
- ☐ Others

(Multiple Choice)

26a) Specify Others

__________________________________

27) Do you consider vaccination an important action to prevent the spread of coronavirus disease?

- ◯ Yes
- ◯ No

28) If a coronavirus disease vaccine were available today, would you agree to be vaccinated with it??

- ◯ Yes
- ◯ No

29) Would you agree to take part in coronavirus disease vaccine trial wherein the trial vaccine would be tested on you?

- ◯ Yes
- ◯ No

29a) Yes. Why?

__________________________________

29b) Why not?

__________________________________

30) How do you think coronavirus disease can be treated

__________________________________

**PART 2: COVID-19 Health Care Behaviour**

**I would now like to ask you some questions about the way you have been seeking care for yourself during this coronavirus pandemic**

31) How did you come to be quarantined?

- ◯ Came in via air travel
- ◯ Came in via land travel
- ◯ Contact tracing by MoH
- ◯ Directed by the national coronavirus helpline, 1025
- ◯ Other

(Choose One)

31a) Specify Others

__________________________________

32) Where were you quarantined?

__________________________________

33) What month(s) were you in quarantine?

◯ March
◯ April
◯ May
◯ June
◯ July
◯ Other
(Choose One)

33a) Specify Others

______________________________
(e.g. capture how many times)

34) How many days were you in quarantine?

◯ 14 days
◯ 21 days
◯ 28 days
◯ Other
(Choose One)

34a) Specify Others

______________________________

35) What do you understand to be the purpose of you having been quarantined

☐ Separating me from community
☐ Monitoring me for signs and symptoms of coronavirus disease
☐ Testing me for coronavirus disease
☐ Protecting me from coronavirus disease
☐ Political reasons
☐ Other
(Multiple Choice)

35a) Specify Others

______________________________

**Of each of the measures I will list, tell me which ones you did or did not do during quarantine by answering either "Yes" or "No"**

36) Staying at the place you were being quarantined at every day

◯ Yes
◯ No

37) Not allowing any visitors

◯ Yes
◯ No

38) Separating yourself from others at the place you were being quarantined

◯ Yes
◯ No

39) Not sharing food and utensils with others

◯ Yes
◯ No

40) Wearing a face mask if you had to leave your room

◯ Yes
◯ No

41) What do you think about your overall quarantine experience?

______________________________

42) What do you think are the benefits of quarantine?

______________________________

43) What do you think are the disadvantages of quarantine?

__________________________________

44) How do you think the Ministry of Health could improve the experience of quarantined individuals?

__________________________________

44a) Specify Others

__________________________________

45) What would you do FIRST if you experience symptoms of coronavirus disease today?

- ◯ I will call the national coronavirus helpline on 1025
- ◯ I will look for someone to advise me on what to do
- ◯ I will go to the hospital / health facility
- ◯ I will go to buy medicines at the pharmacy
- ◯ I will drink local herbal remedies
- ◯ I will quarantine myself at home
- ◯ Other

(Choose One)

45a) Specify Others

__________________________________

**PART 3: COVID-19 Related Stigma**

**Now, I would like to ask you some questions about any judgement/unfairness/shame/blame you may have felt or experienced before, during and after quarantine**

**Did you experience any of these before going into quarantine?**

| | Yes | No |
|---|---|---|
| 46) People kept suggesting you go into quarantine | ◯ | ◯ |
| 47) Family and friends refusing to visit you at home | ◯ | ◯ |
| 48) Being called names to make you feel ashamed | ◯ | ◯ |
| 49) Feeling ashamed because of how you were taken into quarantine | ◯ | ◯ |
| 50) Other | ◯ | ◯ |

50a) Specify Others

__________________________________

#### Did you experience any of these while you were in quarantine

| | Yes | No |
|---|---|---|
| 51) People kept gossiping about you | ◯ | ◯ |
| 52) People keeping their distance (out of fear) | ◯ | ◯ |
| 53) People had a judgmental attitude | ◯ | ◯ |
| 54) Anyone who called thought you were being treated for coronavirus disease | ◯ | ◯ |
| 55) Other | ◯ | ◯ |

55a) Specify Others ______________________________

#### Did you experience any of these after completing quarantine

| | Yes | No |
|---|---|---|
| 56) People kept gossiping about you | ◯ | ◯ |
| 57) Family and friends refused to visit you at home | ◯ | ◯ |
| 58) Anyone who called thought you had been treated for coronavirus disease | ◯ | ◯ |
| 59) Being called names to make you feel ashamed | ◯ | ◯ |
| 60) Other | ◯ | ◯ |

60a) Specify Others ______________________________

61) Did you experience any of these effects after completing quarantine?

- ☐ Shame
- ☐ Self-blame
- ☐ Loneliness
- ☐ Stress
- ☐ Anger
- ☐ Withdrawal from daily activities
- ☐ Strained relationship with family
- ☐ Other

(Multiple Choice)

61a) Specify Others ______________________________

62) Which groups of people do you think the coronavirus disease is generating judgement/unfairness/shame/blame against in your community

☐ Anyone who called the national coronavirus helpline 1025
☐ Anyone who is seen coughing or sneezing in public
☐ Anyone who recovered from coronavirus disease and returned to the community
☐ Family members of people who recovered from coronavirus disease
☐ Anyone who was put into quarantine by the government and returned to the community
☐ Family members of people who were quarantined by the government
☐ Anyone who came into The Gambia by air or land
☐ family members of people came into The Gambia by air or land
☐ Other
(Multiple Choice)

62a) Specify Others

__________________________________

63) What are your feelings towards people diagnosed with coronavirus disease?

☐ They are unfortunate people (bad luck)
☐ They brought the disease upon themselves by travelling during a pandemic
☐ They should be isolated from their family, friends and community until fully recovered
☐ They are people like everyone else who just happened to contract a disease
☐ Their condition was meant to happen (divine will of God)
☐ They did something bad (sinful or immoral behaviour) that is why they have the disease
☐ They should not be allowed to resume work until fully recovered
☐ Other
(Multiple Choice)

63a) Specify Others

__________________________________

**Answer how much you agree or disagree with the statements I will now read out to you.**

| | Strongly Agree | Agree | Disagree | Strongly Disagree |
|---|---|---|---|---|
| 64) People who recover from coronavirus disease will be shamed and blamed for having it | ◯ | ◯ | ◯ | ◯ |
| 65) Public judgement/shaming is caused by their fear of being infected with coronavirus disease | ◯ | ◯ | ◯ | ◯ |
| 66) People may refuse to go into quarantine for fear of being accused of having coronavirus disease | ◯ | ◯ | ◯ | ◯ |
| 67) Recovering from coronavirus disease is nothing to be ashamed about or blamed for | ◯ | ◯ | ◯ | ◯ |
| 68) Judgement/shaming is hurtful and can negatively affect the mental health of the person | ◯ | ◯ | ◯ | ◯ |

**PART 4: Demographics**

**Lastly, I would like to ask you some background questions such as your age group and ethnicity**

69) Age group

◯ 18 - 29
◯ 30 - 39
◯ 40 - 49
◯ 50 - 59
◯ 60 - 69
◯ 70 - 79
◯ 80+

70) Nationality

◯ Gambian
◯ Non-Gambian

70a) Specify Other Nationality

________________________________

71) Marital status

◯ Married
◯ Not married

72) Ethnicity

◯ Mandinka
◯ Fula
◯ Wolof
◯ Other
(Choose One)

72a) Specify Others

________________________________

73) Religion

◯ Islam
◯ Christianity
◯ Other
(Choose One)

73a) Specify Others

__________________________________

74) Education

◯ None
◯ Primary (Grades 1-6)
◯ Junior Secondary (Grades 7-9)
◯ Higher secondary (Grades 10-12)
◯ Tertiary (University education)
◯ Other
(Choose One)

74a) Specify Others

__________________________________

75) Main Occupation

__________________________________
(Please specify)

76) End time

__________________________________
